# Optimized Feature Selection and Advanced Machine Learning for Stroke Risk Prediction in Revascularized Coronary Artery Disease Patients

**DOI:** 10.1101/2025.03.14.25324005

**Authors:** Yong Si, Armin Abdollahi, Negin Ashrafi, Greg Placencia, Elham Pishgar, Kamiar Alaei, Maryam Pishgar

## Abstract

**Background:** Coronary artery disease (CAD) is a leading cause of mortality, with stroke being a major complication following coronary revascularization procedures such as percutaneous coronary intervention (PCI) and coronary artery bypass grafting (CABG). While machine learning (ML) has been used to predict postoperative outcomes, a gap remains in quantifying stroke risk in revascularized CAD patients. This study aims to develop and validate ML models for stroke risk prediction, enhancing clinical decision-making.

**Methods:** We extracted 5,757 patients from the Medical Information Mart for Intensive Care IV (MIMIC-IV) database and applied Pearson correlation analysis, least absolute shrinkage and selection operator (LASSO), ridge regression, and elastic net for feature selection. The initial 35 features were reduced to 14. The dataset was split into training (70%), testing (15%), and validation (15%). We evaluated multiple ML models, including logistic regression, XGBoost, random forest, AdaBoost, Bernoulli naive Bayes, k-nearest neighbors (KNN), and CatBoost. Model performance was assessed using the area under the receiver operating characteristic curve (AUC-ROC) and 500 bootstrapped 95% confidence intervals (CIs).

**Results:** CatBoost achieved the highest performance with an AUC of 0.8486 (95% CI: 0.81240.8797) on the test set and 0.8511 (95% CI: 0.82030.8793) on validation. Shapley Additive Explanations (SHAP) identified Charlson Comorbidity Index (CCI), length of stay (LOS), and treatment types as key predictors. Our model outperformed existing studies, improving AUC by 9% while using a more refined feature set.

**Conclusions:** Integrating multiple feature selection methods, our streamlined model improves efficiency and reliability. The proposed CatBoost model offers a high-accuracy approach to predicting postoperative stroke in CAD patients undergoing revascularization, supporting clinical decision-making and preventive strategies.

## 1 Introduction

Coronary Artery Disease, abbreviated as CAD, is characterized by the narrowing or blockage of the coronary arteries due to atherosclerosis, and it significantly contributes to the high mortality rates associated with cardiovascular diseases [1]. In 2022, heart disease remained the leading cause of death in the U.S., with 702,880 fatalities, equating to one in every five deaths. Coronary heart disease specifically was responsible for 371,506 deaths that year [2]. Revascularization procedures such as PCI and CABG are common and effective treatments for managing CAD [3]. While these revascularization procedures are endorsed by reducing symptoms of CAD, they have limitations and carry unignorable risks. A study from Finland reported that patients with a history of stroke were less than half as likely to receive PCI. Similarly, the presence of Dementia or Alzheimers disease reduced PCI use to the same degree as being over 85 years old [4, 5]. CABG also brings new problems. For instance, another study found that within 10 to 15 years after CABG, up to 40% of patients may require redo CABG due to recurrent angina, late myocardial infarction, and the need for additional intervention, leading to increased risk and cost [6, 7]. Stroke is also a significant risk factor following revascularization. Patients who suffered a stroke within 30 days of the procedure had a substantially higher five-year mortality rate [8, 9, 10, 11]. The risk of stroke within the first year after revascularization was five times higher than in the age- and sex-matched general population. This risk was particularly elevated in patients with a history of stroke, diabetes mellitus, advanced age, male sex, and low socioeconomic status [12, 13]. The prediction of such adverse postoperative events is crucial for improving clinical decision-making and patient care.

Predicting postoperative stroke risk in revascularized CAD patients is crucial for improving clinical decision-making and patient outcomes. Recent studies have demonstrated the potential of machine learning models in predicting in-hospital mortality for ICU patients with heart failure, highlighting the impact of advanced feature selection and model optimization on predictive accuracy [14, 15]. However, research specifically linking stroke risk to revascularization procedures using ML remains limited. Existing studies often face challenges such as overfitting, inadequate feature selection, and class imbalances [16, 17, 18, 19].

This research focuses on optimizing feature selection and enhancing machine learning model performance to predict the risk of postoperative stroke in CAD patients undergoing revascularization. The study aims to advance feature selection techniques, improve predictive accuracy, refine data preprocessing strategies, and enhance model explainability and generalizability. A sophisticated feature selection methodology was employed, integrating four robust statistical techniques: Pearson Correlation Analysis, Preprint – Optimized Feature Selection and Advanced Machine Learning for Stroke Risk Prediction in Revascularized Coronary Artery Disease Patients

LASSO Regression, Ridge Regression, and Elastic Net Regression. This approach reduced the feature set from 35 variables to 14, retaining only the most statistically and clinically relevant predictors. By minimizing redundancy and reducing computational complexity, the streamlined feature set significantly improved model efficiency without compromising accuracy. This rigorous process also mitigates overfitting, underscoring the critical role of high-impact predictor selection in developing robust machine learning models for clinical applications.

The proposed model demonstrated exceptional predictive performance, with the CatBoost algorithm achieving an AUC of 0.8486 (95% CI: 0.81240.8797) on the test set and 0.8511 (95% CI: 0.82030.8793) on the validation set. These results surpass those reported in prior studies, highlighting the effectiveness of the feature selection and preprocessing strategies employed. Consistent and high-performance metrics across datasets establish the CatBoost model as a reliable tool for predicting postoperative stroke risk in CAD patients.

The research underscores the pivotal role of data preprocessing in enhancing model accuracy and reliability. Missing values were addressed through random forest interpolation, maintaining data integrity for both categorical and numerical variables. To address the class imbalance between stroke and non-stroke cases, the study applied the Synthetic Minority Over-sampling Technique (SMOTE), which improved the model’s sensitivity to rare events. Prior work has shown that SMOTE can effectively enhance model performance in clinical settings by generating synthetic minority class samples and reducing bias in predictive analytics [20]. Additionally, categorical features were optimized through feature category reduction, mitigating overfitting and improving computational efficiency. These preprocessing steps collectively contributed to the models robustness and ensured its applicability in diverse clinical scenarios.

To improve model interpretability, SHAP analysis was incorporated, elucidating the contribution of individual predictors to the CatBoost model. The CCI emerged as the most significant variable, followed by LOS and treatment type. By visualizing the impact of these predictors, the study enhances the model’s transparency, making it a valuable decision-support tool for clinicians. The integration of SHAP analysis bridges the gap between computational outputs and clinical applicability, ensuring predictions are both interpretable and actionable.

The study also emphasizes the importance of generalizability across diverse clinical settings. Prior research on mortality prediction in CAD patients has demonstrated the value of integrating machine learning models with clinical decision support systems to enhance patient risk assessment [21]. While SMOTE effectively balanced the dataset, we acknowledge its potential to alter class proportions and advocate for further validation using external datasets. This focus on generalizability reflects a commitment to developing predictive tools that are not only accurate but also practical for real-world implementation. Collectively, these contributions advance the field of predictive analytics, offering a comprehensive framework for risk stratification and personalized care in CAD management.

## 2 Methodology

### 2.1 Data Source

The MIMIC-IV database is a publicly accessible, comprehensive dataset that provides detailed health-related information on patients admitted to critical care units at the Beth Israel Deaconess Medical Center in Boston, Massachusetts [22]. Building upon its predecessor, MIMIC-III, MIMIC-IV offers an extensive collection of data, including patient admissions, demographics, vital signs, laboratory results, procedures, medications, caregiver notes, imaging reports, and detailed mortality information, such as dates and times. This robust dataset is specifically designed to support research in critical care, facilitating the development and validation of predictive models and enabling clinical studies aimed at enhancing patient outcomes. The breadth and depth of MIMIC-IV’s records establish it as an invaluable resource for researchers investigating various dimensions of intensive care medicine.

### 2.2 Data Extraction

Our study focused on adult intensive care unit (ICU) patients diagnosed with CAD who underwent revascularization procedures. Using text matching of ICD-9 and ICD-10 codes, we initially identified a cohort of 5,988 patients who had undergone either PCI or CABG. Patients under 18 years of age or with ICU stays of less than 24 hours were excluded, resulting in a final cohort of 5,757 adult patients with common clinical presentations. This refined dataset was designed to provide meaningful insights into the management and outcomes of CAD patients undergoing revascularization within critical care settings. The data extraction process is outlined in Figure 1.

**Figure 1:**
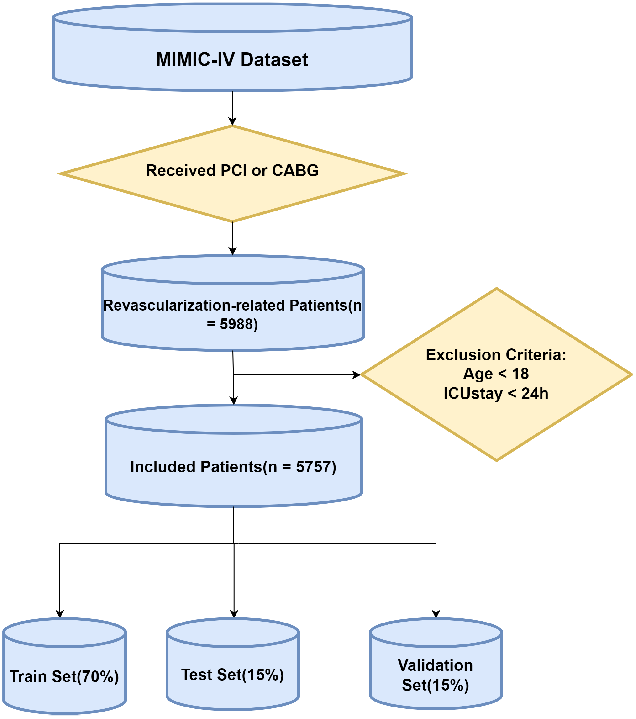
Flowchart of the data extraction procedure.

### 2.3 Feature Extraction and Feature Selection

Through a literature review and expert consultation, we identified and selected key features for analysis following data extraction. From a cohort of 5,757 patients, a comprehensive set of 35 features was curated, encompassing demographic, clinical, physiological, and treatment-related variables. Demographic variables included age, gender, race, insurance type, marital status, and the initial care unit (categorized as Cardiac Care Unit [CCU], Cardiovascular Intensive Care Unit [CVICU], or others). Clinical and physiological variables comprised the Charlson Comorbidity Index (CCI), Glasgow Coma Scale (GCS) score, and vital signs, including, systolic and diastolic blood pressure, temperature, heart rate, respiratory rate, and oxygen saturation. Additionally, a broad range of laboratory parameters were considered, such as hemoglobin, red blood cell distribution width, white blood cell count, platelet count, hematocrit, creatinine, prothrombin time (PT), international normalized ratio (INR), partial thromboplastin time (PTT), blood urea nitrogen (BUN), glucose, calcium, sodium, chloride, bicarbonate, and lactate levels.

Treatment-related variables included revascularization modalities (CABG and PCI), history of stroke, antiplatelet usage, and LOS. The primary outcome, postoperative stroke, was defined as a binary variable (Yes/No). Pre-existing features were directly extracted from the MIMIC-IV database, while additional variables were derived using ICD codes and item IDs. This robust feature set was designed to ensure a comprehensive evaluation of factors influencing postoperative stroke outcomes in this patient population.

For feature selection, we employed four advanced methodologies: Correlation Analysis, Lasso Regression, Ridge Regression, and Elastic Net. Each technique offered unique insights into feature importance, enabling a robust cross-validation process to identify the most predictive variables. Figures 2, 3, 4, and 5 depict the feature rankings derived from these methods. Following this rigorous selection process, a final set of 14 features was identified. The selected demographic variables included anchor age, race, insurance status, the CCI score, and a history of stroke. Vital signs, such as systolic blood pressure (SBP) and respiratory rate, were incorporated, alongside laboratory markers, including calcium, sodium, glucose, hematocrit, and international normalized ratio (INR). LOS were also included. These features were chosen based on consistently high importance scores across all four methodologies, ensuring their relevance and predictive strength in modeling the risk of postoperative stroke among revascularized CAD patients.

**Figure 2:**
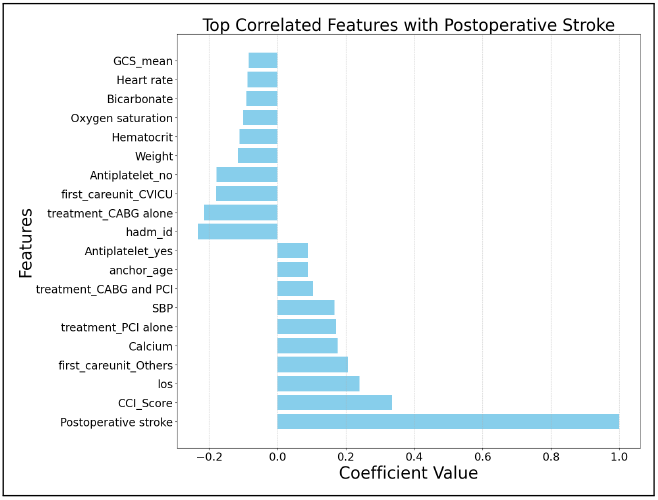
Feature ranking from correlation analysis.

**Figure 3:**
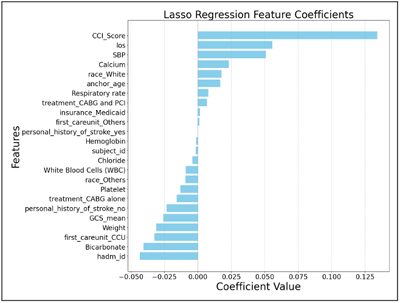
Feature ranking from Lasso regression analysis.

**Figure 4:**
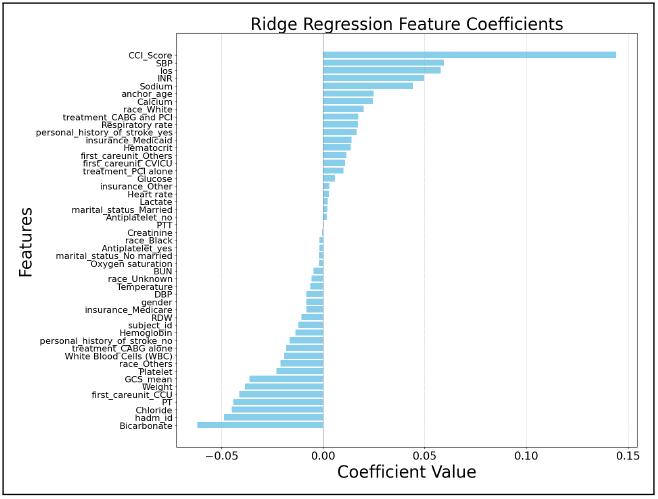
Feature ranking from Ridge regression analysis.

**Figure 5:**
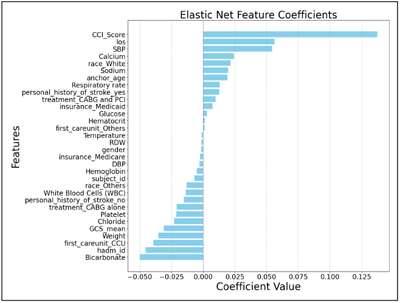
Feature ranking from elastic net feature coefficients analysis.

### 2.4 Data Preprocessing

The preprocessing of the dataset involved a comprehensive and methodical approach to ensure data integrity and enhance the quality of input for subsequent analysis. Missing values were addressed using a dual imputation strategy: Regular Random interpolation was applied for categorical variables, while Random Forest-based interpolation was utilized for numerical variables. This combined approach ensured both consistency and accuracy across the dataset.

To further refine the dataset, Feature Category Reduction was performed to consolidate categories within categorical variables, mitigating the risk of overfitting and enhancing computational efficiency. Imbalances in the dataset, particularly in the target variable (postoperative stroke) and certain feature variables, were addressed using the SMOTE. SOMTE is advantageous due to its simplicity and effectiveness in addressing class imbalance by generating synthetic samples rather than merely replicating existing ones. This enhances the classifier’s generalization ability, reducing the risk of overfitting associated with traditional oversampling methods [23]. SMOTE effectively balanced the dataset by generating synthetic examples of the minority class, ensuring equitable representation and facilitating the detection of rare but clinically significant events. Categorical variables were encoded into numerical formats using label encoding, enabling seamless integration into feature selection models.

The training dataset exhibited a pronounced class imbalance, with significantly fewer postoperative stroke cases compared to non-stroke cases. SMOTE was applied to oversample the minority class, creating synthetic samples to balance the class distribution. This preprocessing step enabled the models to better identify patterns within the minority class, thereby improving predictive performance and ensuring fairness across variables.

Through these preprocessing techniques, the dataset was meticulously prepared, enhancing its suitability for feature selection and modeling while improving the ability to detect critical but infrequent outcomes.

### 2.5 Model Training and Hyperparameter Tuning

The dataset was partitioned into training (70%), validation (15%), and test (15%) sets to facilitate robust model evaluation. Standardization was applied to all numerical features, ensuring data normalizationa critical step for machine learning algorithms sensitive to input scale, such as Support Vector Machines (SVM) and Logistic Regression.

A suite of machine learning models was evaluated for predicting postoperative stroke in a clinical dataset. The models included Logistic Regression (LR), Random Forest (RF), SVM, XGBoost, AdaBoost, Bernoulli Naïve Bayes (NB), and KNN. Each model underwent extensive hyperparameter tuning using GridSearchCV with cross-validation to identify optimal configurations. For instance, LR achieved its best performance with an L1 penalty and C=0.001, RF with max_depth=2 and n_estimators=15, and SVM with a linear kernel and C=0.1. Similarly, XGBoost and AdaBoost were optimized for hyperparameters such as maximum depth, learning rate, and ensemble size, while KNN employed dimensionality reduction via Principal Component Analysis (PCA). Although these models demonstrated strong individual performance, inconsistencies were observed across evaluation metrics.

Among the models, the CatBoost algorithm emerged as the most effective due to its inherent ability to process categorical variables without prior encoding and its robustness when handling highly imbalanced datasets. Comprehensive hyperparameter tuning was performed using GridSearchCV, optimizing parameters such as iterations ([5, 10, 20]), learning rate ([0.5, 0.7, 1.0]), depth ([1, 2]), L2 regularization ([500, 1000, 2000]), bagging temperature ([5, 10, 15]), random strength ([10, 30, 50]), and feature sampling (RSM: [0.1, 0.2, 0.3]).

The final CatBoost model configuration20 iterations, a learning rate of 1.0, depth of 2, L2 regularization (l2_leaf_reg) of 500, and bagging temperature of 5achieved superior performance. It recorded the highest cross-validated AUC score of 0.8933 on the training set, with strong generalizability reflected by AUC scores of 0.8486 and 0.8511 on the test and validation sets, respectively. This consistency across datasets highlights the robustness and predictive power of the optimized CatBoost model for identifying postoperative stroke risk in revascularized CAD patients.

### 2.6 Statistical Analysis

We conducted paired t-tests, chi-square tests, and calibration analyses to assess the performance and reliability of the models across training and validation datasets. A threshold p-value of 0.05 was applied to evaluate the statistical significance of differences. The paired t-test results indicated no statistically significant difference between the training and validation AUC scores, confirming the consistency and generalizability of the models. However, chi-square tests revealed a significant difference in feature distributions between the training and validation sets, likely attributable to the SMOTE used to address class imbalance. SMOTE effectively balances datasets by generating synthetic examples for minority classes but can inadvertently alter the original proportions of categorical variables, particularly class labels, thereby impacting the distribution.

Calibration tests were also employed to evaluate the alignment of predicted probabilities with actual event frequencies, a critical aspect of model reliability in clinical applications. Calibration ensures that a models predicted probabilities correspond to observed outcomes, enabling actionable and trustworthy risk assessments [24]. While discrimination metrics assess a models ability to differentiate between outcomes, poor calibration can undermine the reliability of probability estimates, limiting their practical utility in clinical decision-making.

The calibration analyses of the promoted CatBoost model demonstrated its robustness for clinical applications. Calibration curves showed consistent alignment between predicted probabilities and observed outcomes across both training and validation datasets. This finding indicates that the CatBoost model is well-calibrated, providing reliable probability estimates for postoperative stroke risk. Such calibration reinforces the models practical utility, ensuring clinicians can confidently use its outputs for informed decision-making in high-stakes scenarios. These results highlight the importance of combining discrimination and calibration assessments to develop predictive models that are both accurate and clinically actionable.

### 2.7 Feature Impact

We employed SHAP to evaluate and interpret feature impacts within the predictive model. SHAP is a powerful tool in machine learning, particularly valuable in clinical research, as it provides interpretable and actionable insights into model predictions [25]. By assigning Shapley values to each feature, SHAP quantifies the individual contribution of features to specific predictions, offering a transparent view of the decision-making processes of complex models. This transparency is especially critical in clinical settings, where trust and interpretability are essential for the effective application of predictive models.

In the context of predicting postoperative stroke, SHAP analysis enabled us to uncover the influence of key factors such as patient age, comorbidities, vital signs, and treatment types on the model’s risk assessments. By elucidating these contributions, SHAP fosters confidence in the model’s predictions and supports clinicians in identifying modifiable risk factors, which can inform targeted clinical interventions. Furthermore, SHAP values are instrumental in detecting potential biases or anomalies within the model by highlighting unexpected feature contributions, facilitating necessary adjustments to improve model fairness and accuracy.

In conclusion, SHAP analysis serves as an indispensable component of clinical machine learning by bridging the gap between complex predictive models and the imperative for interpretability in medical decision-making. Through detailed decomposition of feature contributions, SHAP empowers clinicians to make data-driven, informed decisions, ultimately enhancing patient outcomes and fostering trust in AI-driven healthcare solutions.

## 3 Results

### 3.1 Evaluation Results

The performance of each predictive model was rigorously evaluated using a comprehensive suite of metrics, including the AUC, Receiver Operating Characteristic (ROC) AUC, Precision-Recall Curve, Sensitivity, Specificity, Average Precision, F1 Score, Confusion Matrix, and Log-Loss. This multifaceted evaluation framework provided a holistic understanding of each models ability to accurately predict postoperative stroke which is shown in Table 1.

**Table 1:**
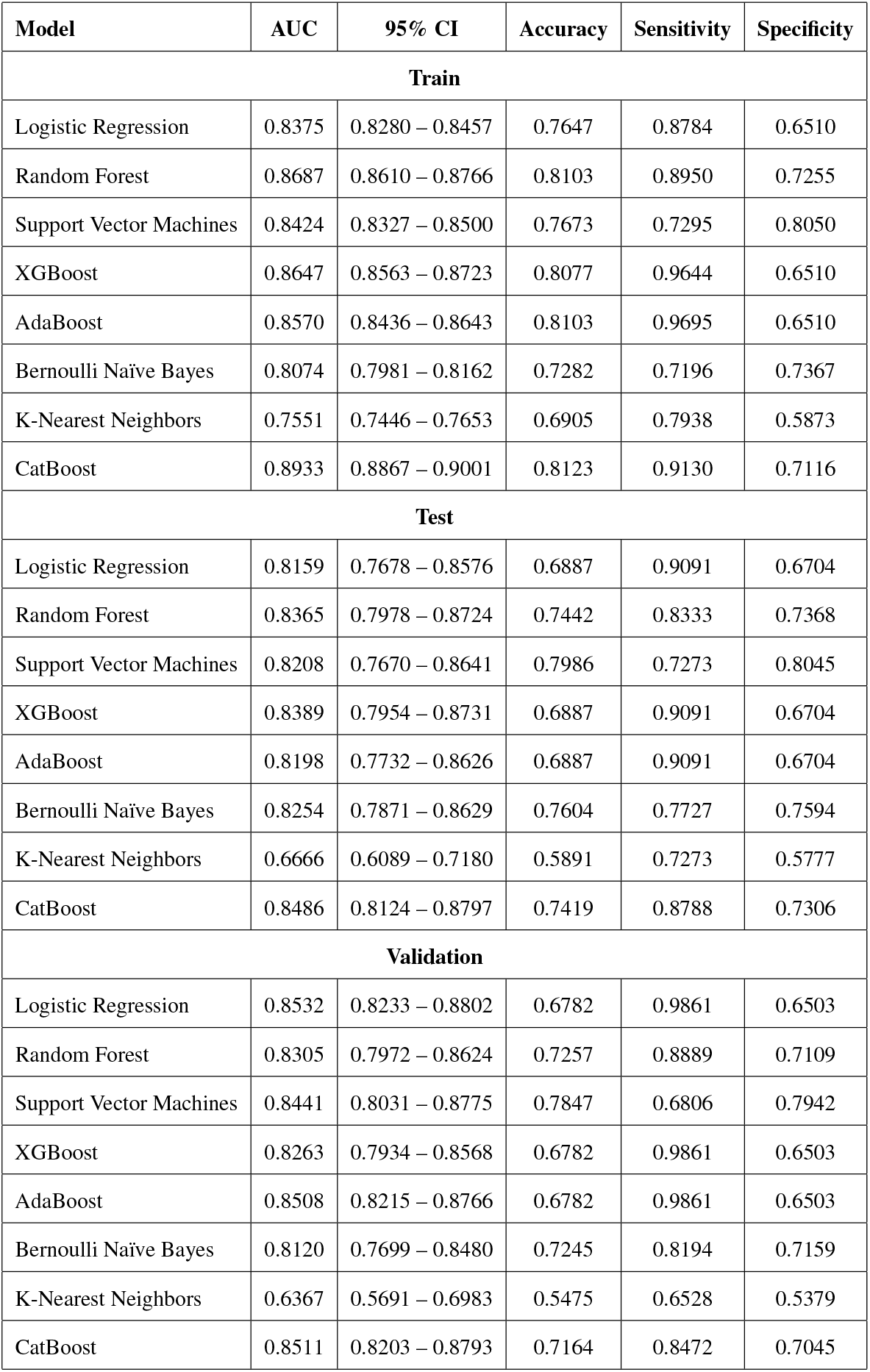
Performance Metrics of Models Built.

| Model | AUC | 95% CI | Accuracy | Sensitivity | Specificity |
| --- | --- | --- | --- | --- | --- |
| **Train** |  |  |  |  |  |
| Logistic Regression | 0.8375 | 0.8280 – 0.8457 | 0.7647 | 0.8784 | 0.6510 |
| Random Forest | 0.8687 | 0.8610 – 0.8766 | 0.8103 | 0.8950 | 0.7255 |
| Support Vector Machines | 0.8424 | 0.8327 – 0.8500 | 0.7673 | 0.7295 | 0.8050 |
| XGBoost | 0.8647 | 0.8563 – 0.8723 | 0.8077 | 0.9644 | 0.6510 |
| AdaBoost | 0.8570 | 0.8436 – 0.8643 | 0.8103 | 0.9695 | 0.6510 |
| Bernoulli Naïve Bayes | 0.8074 | 0.7981 – 0.8162 | 0.7282 | 0.7196 | 0.7367 |
| K-Nearest Neighbors | 0.7551 | 0.7446 – 0.7653 | 0.6905 | 0.7938 | 0.5873 |
| CatBoost | 0.8933 | 0.8867 – 0.9001 | 0.8123 | 0.9130 | 0.7116 |
| **Test** |  |  |  |  |  |
| Logistic Regression | 0.8159 | 0.7678 – 0.8576 | 0.6887 | 0.9091 | 0.6704 |
| Random Forest | 0.8365 | 0.7978 – 0.8724 | 0.7442 | 0.8333 | 0.7368 |
| Support Vector Machines | 0.8208 | 0.7670 – 0.8641 | 0.7986 | 0.7273 | 0.8045 |
| XGBoost | 0.8389 | 0.7954 – 0.8731 | 0.6887 | 0.9091 | 0.6704 |
| AdaBoost | 0.8198 | 0.7732 – 0.8626 | 0.6887 | 0.9091 | 0.6704 |
| Bernoulli Naïve Bayes | 0.8254 | 0.7871 – 0.8629 | 0.7604 | 0.7727 | 0.7594 |
| K-Nearest Neighbors | 0.6666 | 0.6089 – 0.7180 | 0.5891 | 0.7273 | 0.5777 |
| CatBoost | 0.8486 | 0.8124 – 0.8797 | 0.7419 | 0.8788 | 0.7306 |
| **Validation** |  |  |  |  |  |
| Logistic Regression | 0.8532 | 0.8233 – 0.8802 | 0.6782 | 0.9861 | 0.6503 |
| Random Forest | 0.8305 | 0.7972 – 0.8624 | 0.7257 | 0.8889 | 0.7109 |
| Support Vector Machines | 0.8441 | 0.8031 – 0.8775 | 0.7847 | 0.6806 | 0.7942 |
| XGBoost | 0.8263 | 0.7934 – 0.8568 | 0.6782 | 0.9861 | 0.6503 |
| AdaBoost | 0.8508 | 0.8215 – 0.8766 | 0.6782 | 0.9861 | 0.6503 |
| Bernoulli Naïve Bayes | 0.8120 | 0.7699 – 0.8480 | 0.7245 | 0.8194 | 0.7159 |
| K-Nearest Neighbors | 0.6367 | 0.5691 – 0.6983 | 0.5475 | 0.6528 | 0.5379 |
| CatBoost | 0.8511 | 0.8203 – 0.8793 | 0.7164 | 0.8472 | 0.7045 |

Among the models, the CatBoost algorithm consistently outperformed its counterparts, achieving the highest AUC scores across all datasets: 0.8486 on the test set and 0.8511 on the validation set. These results underscore CatBoost’s superior capability to discriminate between positive and negative classes. Furthermore, the model demonstrated a balanced performance with a sensitivity of 87.88% and a specificity of 73.06%, effectively minimizing false positive and false negative rates. Figures 6, 7, and 8 present the ROC curves for all models on the training, test, and validation sets, respectively.

**Figure 6:**
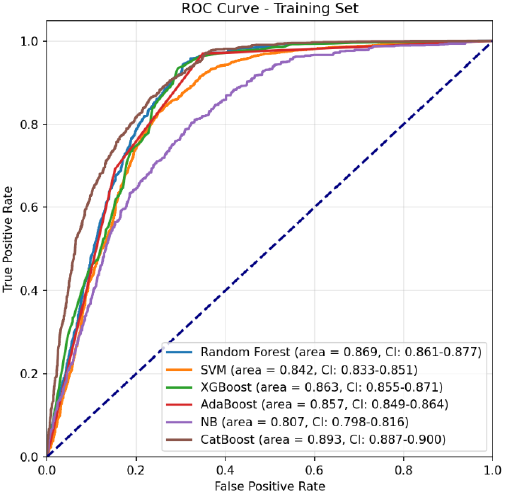
ROC Curve (Train Set).

**Figure 7:**
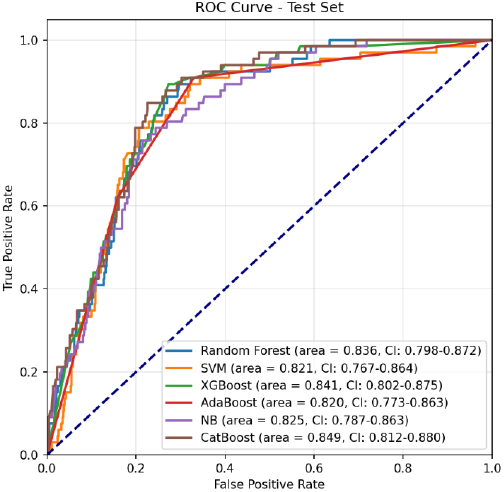
ROC Curve (Test Set).

**Figure 8:**
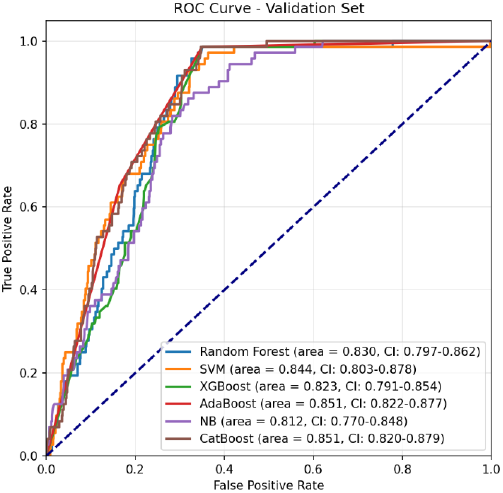
ROC Curve (Validation Set).

Calibration testing in Figure 9 further validated CatBoosts reliability in probabilistic predictions. The model achieved a Brier score loss of 0.130 on the training set and 0.160 on the validation set, indicating robust probability estimation with minimal uncertainty. While slight discrepancies in calibration

**Figure 9:**
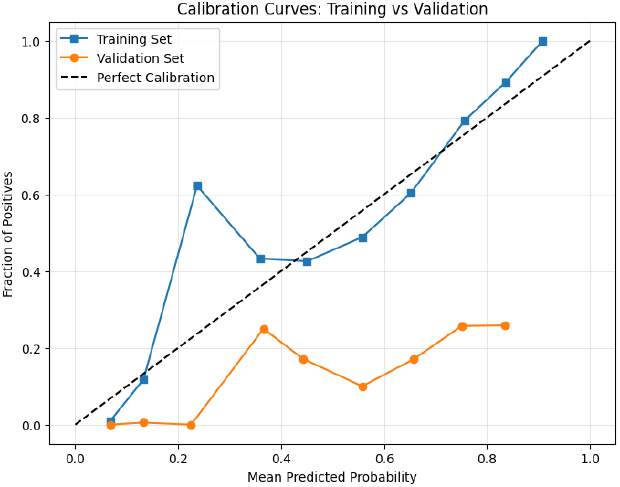
Calibration Curves: Training vs. Validation.

Although Random Forest and XGBoost models delivered competitive performance, CatBoost distinguished itself by seamlessly handling categorical data, maintaining consistent stability across datasets, and offering superior interpretability through SHAP analysis. The models ability to directly process categorical features without additional preprocessing steps further highlights its utility in clinical datasets.

In summary, the CatBoost model excelled not only in discriminatory power, as evidenced by its high AUC scores, but also in generating well-calibrated probability estimates. These findings underscore CatBoost’s potential for clinical deployment, where accurate, interpretable, and reliable predictions are vital for risk stratification and informed decision-making.

### 3.2 SHAP Results

To evaluate the contribution of individual variables to the predictive model, we employed SHAP analysis. SHAP values provide interpretable insights into model predictions, offering transparency in understanding the rationale behind diagnostic decisions. This interpretability is particularly valuable in healthcare, as it enables clinicians to comprehend and trust the factors driving the model’s predictions.

The analysis, depicted in Figure10 and Figure11 includes a ranked visualization of variable importance, presented in the figure below, which highlights the mean absolute SHAP values for each feature. This bar plot quantifies the average magnitude of each features impact on the models predictions, providing a clear overview of their relative contributions. Among the evaluated features, the CCI score emerged as the most influential variable, followed by LOS and treatment types. These three factors were identified as critical determinants of postoperative stroke risk in revascularized CAD patients.

**Figure 10:**
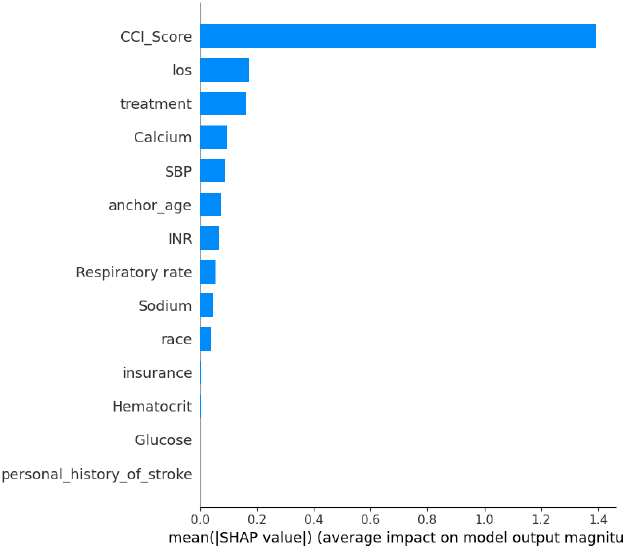
SHAP analysis of top features.

**Figure 11:**
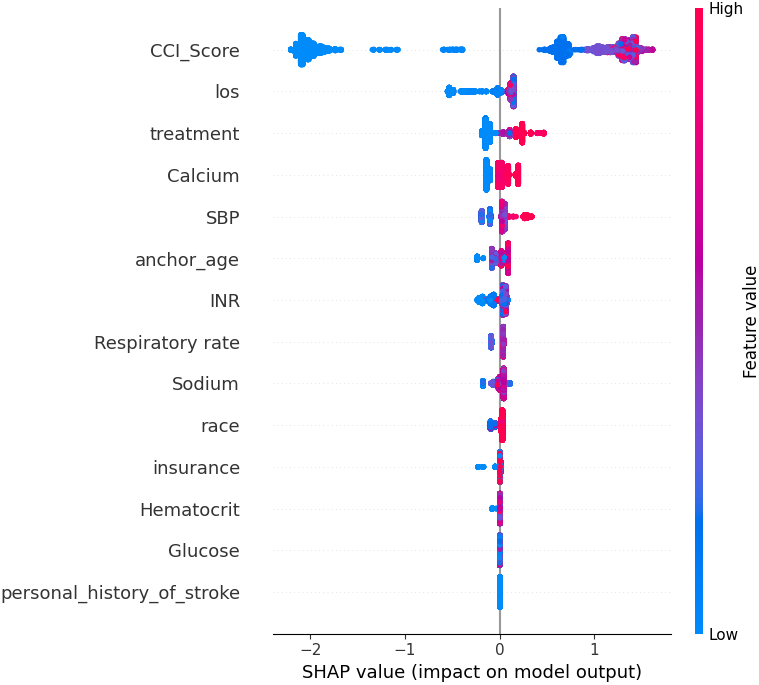
SHAP analysis of top features.

By prioritizing these variables, SHAP analysis underscores their significance in risk stratification, offering actionable insights for clinical decision-making and potential areas for intervention.

## 4 Discussion

This study aims to predict the risk of postoperative stroke in CAD patients undergoing revascularization procedures by leveraging optimized feature selection and ML modeling. Our findings demonstrate significant improvements in predictive accuracy and clinical applicability compared to existing methodologies.

A benchmark study conducted by Lin et al. (2024) [13] explored stroke prediction in revascularized CAD patients using ML models, including CatBoost, logistic regression, and SVM. Their CatBoost model achieved an AUC of 0.760 on the test set, identifying the CCI as the most important predictor. While their study underscored the potential of ML in this domain, it had notable limitations: (a) reliance on an extensive feature set increased the risk of overfitting and reduced model interpretability; (b) inadequate handling of class imbalances and missing data likely skewed performance metrics; and (c) the absence of external validation limited the generalizability of their findings to broader clinical settings. Our study addresses these limitations through several key advancements:

Optimized Feature Selection: We implemented a multi-method approach integrating Pearson correlation analysis, LASSO, ridge regression, and elastic net regression to refine the initial feature set from 35 to 14 variables. This strategy ensured the inclusion of statistically and clinically relevant predictors, reduced feature redundancy, minimized computational complexity, and enhanced overall model efficiency.

Enhanced Predictive Accuracy: Using the optimized CatBoost model, we achieved an AUC of 0.8486 on the test set, representing a 9% improvement over the performance reported by Lin et al. This enhancement underscores the effectiveness of our rigorous feature selection, data preprocessing, and model optimization techniques.

Increased Model Robustness: By incorporating comprehensive bootstrapping techniques and addressing class imbalances using SMOTE, our model demonstrated improved generalizability and resilience across diverse datasets. Furthermore, the integration of SHAP analysis provided actionable insights into feature importance, facilitating interpretability and bridging the gap between computational predictions and clinical decision-making.

## 5 Limitation

Despite these advancements, our study has certain limitations. The exclusive use of the MIMIC-IV database constrains external validation, limiting the applicability of our findings to broader populations. Future research should incorporate external datasets from diverse institutions to ensure the generalizability of the model. Additionally, while SMOTE effectively addressed class imbalances, it may alter the original class distribution, requiring careful interpretation of results. Finally, translating ML models into routine clinical practice necessitates further exploration of user-friendliness, clinician acceptance, and integration into existing healthcare workflows.

## 6 Conslusion

This study significantly advances stroke risk prediction in revascularized CAD patients through the implementation of optimized feature selection and ML modeling. By employing the CatBoost algorithm, the study achieved superior predictive performance, highlighted by high AUC scores and enhanced interpretability using SHAP analysis. These findings underscore the utility of data-driven approaches in supporting clinical decision-making and improving patient outcomes.

Despite these advancements, there are several opportunities to enhance the robustness, generalizability, and clinical applicability of this work. Future improvements should address the following:

1. External Validation: The reliance on the MIMIC-IV database limits the generalizability of the findings. Future research should incorporate external datasets from diverse populations and healthcare institutions to validate the models applicability in broader clinical contexts.
2. Longitudinal Studies: Expanding the dataset to include longitudinal patient outcomes could improve predictions of long-term risks. This approach would provide valuable insights into the progression of CAD and the efficacy of various interventions over time.
3. Advanced Modeling Techniques: Further studies could explore ensemble learning by combining multiple ML algorithms to enhance predictive accuracy. Additionally, deep learning models, such as recurrent neural networks (RNNs) or transformers, may be utilized to capture temporal dependencies in the data.
4. Enhanced Feature Selection: While this study employed multiple feature selection techniques, integrating domain-specific knowledge with automated methods could refine the feature set further. Incorporating novel biomarkers or genomic data could reveal additional predictors of stroke risk.

By addressing these areas, future studies can enhance the clinical impact of ML models, providing a comprehensive framework for personalized medicine in CAD management. These advancements will not only improve patient care but also lay the groundwork for broader applications of artificial intelligence in cardiovascular health.

## Data Availability

All data used in this study are publicly available and can be accessed at https://physionet.org/content/mimiciv/2.2/.

## Acknowledgements

We sincerely appreciate the efforts behind the development and publication of MIMIC-IV, which has been instrumental in advancing critical care research.

## Authors’ contributions

Y.S., N.A., and A.A. contributed to all aspects of the study, including conceptualization, methodology, data analysis, and manuscript preparation. E.P. and K.A. provided expert opinions and guidance on the study design and clinical implications. M.P. was responsible for project administration and supervision. All authors reviewed and approved the final manuscript.

## Funding

Not applicable.

## Availability of data and materials

The raw dataset is publicly available in the MIMIC-IV Repository

## Ethics approval and consent to participate

The data utilized in this study is sourced from the Medical Information Mart for Intensive Care version IV (MIMIC-IV), a publicly accessible and de-identified database. As such, it did not necessitate informed consent or approval from an Institutional Review Board. All research procedures adhered to relevant guidelines and regulatory standards.

## Consent for publication

Not applicable.

## Competing interests

The authors declare no competing interests.

## Notes

### Competing Interest Statement

The authors have declared no competing interest.

### Funding Statement

This study did not receive any funding

